# Neurotransmitter‑related structural network damage and language performance after stroke

**DOI:** 10.64898/2026.03.10.26348093

**Authors:** Tom Hornberger, Robert Schulz, Philipp J. Koch, Jan F. Feldheim, Paweł P. Wróbel, Götz Thomalla, Tim Magnus, Dorothee Saur, Fanny Quandt, Benedikt M. Frey

## Abstract

**Background:** Aphasia commonly occurs after left-hemispheric stroke, yet substantial inter-individual variability in language outcomes remains insufficiently explained by established clinical systems neuroscience concepts. Emerging evidence suggests that the integrity of specific neurotransmitter systems may influence functional outcomes after stroke. This study examined whether the damage to neurotransmitter-related structural networks is associated with post-stroke language impairment.

**Methods:** Data of 270 patients with left-hemispheric stroke from two openly available cohorts were analyzed: the acute Washington Stroke Cohort and the chronic Aphasia Recovery Cohort. Neurotransmitter-related network damage was quantified by embedding individual stroke lesion masks into normative connectomes weighted by PET-derived density maps of 16 neurotransmitter receptors and transporters. Partial least squares (PLS) regression identified informative predictors of language functioning, followed by linear regression analyses adjusted for age, sex, lesion volume, and time post-stroke.

**Results:** Across both cohorts, PLS analyses converged on a neurochemical profile in which damage to networks related to serotonergic (5-HT_1a_, 5-HT_2a_) and dopaminergic (D1) receptor distributions showed the strongest associations with poorer language performance. Damage to the 5-HT_1a_ and D1-related networks remained significant in fully adjusted models, leading to substantially improved model fit.

**Conclusion:** The disruption of large-scale serotonergic (5-HT_1a_) and dopaminergic (D1) brain networks is associated with language impairment in acute and chronic stroke. Neurotransmitter-related network damage explained additional variability in language performance beyond clinical variables and lesion burden. This work adds a neurochemically informed network perspective to aphasia research and may pave the way for future biological patient stratification to support targeted rehabilitation strategies, such as pharmacological interventions.

## Introduction

Aphasia is a frequent and disabling consequence of stroke, affecting approximately one third of patients in both the acute and chronic phases of recovery^1^. It is defined as an acquired language disorder caused by brain damage, characterized by impairments across multiple components of the language system, including phonological, morphological, semantic, syntactic and pragmatic aspects^2^. Traditionally, the emergence of aphasia has been linked to damage in specific brain regions, particularly classical language areas such as Broca’s and Wernicke’s areas. While lesion size and location are among the strongest predictors of aphasia severity in acute stroke, they only account for a portion of the observed variance in language outcomes in chronic stroke^3^. In addition to lesion characteristics, demographic and clinical factors such as age^4,5^, time post-stroke^4^, overall health status^5^, and damage to critical regions^6^ have been identified as relevant predictors of aphasia severity. Recent studies have additionally highlighted that the disruption and reorganization of large-scale brain networks are also linked to aphasia. Disconnections of white matter tracts and fragmentation of residual network architecture have been associated with chronic aphasia severity independent of lesion size^7^. Multivariate connectome-based analyses have further revealed that the integrity of connections between spared regions, especially those linking parietal with posterior temporal areas, is crucial for preserving speech and language functions^8^. These findings underscore the importance of network mechanisms, alongside lesion characteristics, in evaluating aphasia pathophysiology and recovery trajectories. However, a substantial proportion of the variance in language impairment remains unexplained by these factors, suggesting that additional neurobiological mechanisms may contribute to individual outcomes.

In an effort to gain a deeper understanding of the neurobiological mechanisms of post-stroke deficits, recent studies have started to complement traditional lesion and connectivity analyses with normative maps of neurotransmitter (NT) receptor and transporter distributions^9,10^. This approach enables integration of molecular-level information into system-level analyses, providing a novel framework for linking structural alterations in brain networks to specific neurochemical systems. Notably, alterations in dopaminergic pathways, as inferred from the distribution of dopamine transporters, have recently been associated with poorer functional recovery following stroke^9^. Furthermore, recent evidence indicates that greater lesion load and network disconnections in several neurotransmitter systems have been associated with worse aphasia recovery^11^. Building on this foundation, the present study explores whether this neurochemical approach may similarly inform understanding of network-level vulnerability of aphasia severity in acute and chronic stroke cohorts.

## Methods

The present study is based on secondary analyses of two independent, previously published and openly available datasets of anonymized patients with stroke. Details regarding dataset access and availability are reported in the Data Availability section.

### Washington Stroke Cohort (acute cohort)

The first cohort is based on previously published data by Corbetta et al. as part of the Washington Stroke Cohort^12^ (WSC). It includes imaging and neurobehavioral data of 132 patients with a first symptomatic stroke, and clinical evidence of any neurological impairment^12^. Participants were assessed at 1-2 weeks post-stroke using an extensive neurobehavioral battery and multimodal MRI, including structural (T1/T2, FLAIR), resting-state functional, diffusion-weighted, and perfusion imaging. Detailed original inclusion and exclusion criteria are provided in the Supplementary Material. For the present analysis, a subset of 44 patients was selected from the original dataset based on additional criteria: (1.) a single left-hemispheric supratentorial ischemic stroke, and (2.) complete language battery assessments at the acute stage. The language battery assessed auditory comprehension, speech production, and reading at both the single-word and sentence level, as well as semantic and phonological processing. To derive a single quantitative measure of overall language performance, raw scores from all language subtests at the acute stage were entered into a principal component analysis. The first principal component, which accounted for 75.2% of variance, was extracted and used as the language factor score, reflecting shared variance across comprehension, production, reading, and phonological and semantic processing. For interpretability, patient-specific scores were linearly rescaled to a 0 – 100 range, with zero corresponding to the lowest and 100 to the highest observed value in the dataset. Ethical approval for data acquisition was obtained from the Institutional Review Board of Washington University School of Medicine (WUSM). All procedures were conducted in accordance with the Declaration of Helsinki. Written informed consent was obtained from all participants prior to enrollment^12^.

### Aphasia Recovery Cohort (chronic cohort)

The second patient cohort is based on data from the Aphasia Recovery Cohort^13^ (ARC) at the University of South Carolina, a comprehensive repository designed to support reproducible research in post-stroke aphasia. ARC consolidates data from multiple prior investigations, including studies on anomia treatment, speech entrainment, brain stimulation in aphasia, and the POLAR (Predicting Outcome in Language Rehabilitation in Aphasia) protocol. Importantly, ARC focuses on patients in the chronic phase of stroke recovery. For the present analysis, only baseline assessments were considered, defined as the first available T2-weighted MRI as well as the first available behavioral testing for each patient. All analyses were based on version 1.0.1 of the ARC dataset. Detailed original inclusion and exclusion criteria are provided in the Supplementary Material. In the present analysis, 226 patients were selected from the original dataset based on having (1.) a T2-weighted MRI scan at the baseline assessment, and (2.) a supratentorial stroke location. Language impairment was assessed using the Western Aphasia Battery-Revised (WAB-R)^14^, a standardized and widely used instrument designed to quantify the type and severity of aphasia following stroke. The WAB-R evaluates several domains of language function, including spontaneous speech, auditory comprehension, repetition, and naming, providing a comprehensive profile of aphasia deficits. For the present analysis, the WAB-R aphasia quotient (WAB-AQ) was used as the primary language performance measure, as provided in the ARC dataset. The WAB-AQ is a composite score derived from the core language subtests and ranges from 0 to 100, with lower scores indicating more severe language impairment. The ARC dataset was approved by the Institutional Review Board at the University of South Carolina, which determined that the anonymized data met safe harbor criteria and were exempt from further review (proposal *“OpenNeuro Clinical”* Pro00132576). The analyses presented here were conducted on de-identified data.

### Image processing

For details about imaging and scanner specifications, please refer to the original publications^12,13^. Lesion masks used in the present study were derived from both original datasets. In the WSC dataset, lesions were manually delineated on atlas-transformed (Talairach and Tournoux, 1988) T1-, T2- and FLAIR weighted MRI scans acquired approximately two weeks post-stroke. Segmentation was performed by trained research staff using the Analyse software and was reviewed by board-certified neurologists^15^. All segmentation procedures were conducted blinded to behavioral performances, and a second expert review was performed to refine lesion boundaries^12^. For the present analyses, the original lesion masks were re-rastered to standard Montreal Neurological Institute (MNI) space dimensions, using the FLIRT tool with nearest-neighbor interpolation from FSL (version 6.0.7)^16^, to ensure compatibility with subsequent processing steps. In the ARC dataset, lesions were manually delineated by an expert on the T2-weighted MRI scan from the first visit^13^. For this analysis, T2-weighted images corresponding to the lesion masks were nonlinearly normalized to the MNI standard space using FreeSurfer’s Easyreg tool^17,18^. The resulting transformation was subsequently applied to the corresponding lesion mask using nearest-neighbor interpolation. Lesion volumes for both datasets were calculated in MNI space by summing the number of voxels within the binarized mask and multiplying by the isotropic voxel volume (1 mm^3^). This procedure ensures comparability across participants due to spatial normalization.

### Neurotransmitter-related network damage

To assess the disruption of NT-related structural networks, the binarized lesion masks in MNI space were mapped onto normative NT-related structural connectome data, as described previously by Koch et al.^9^. In summary, the normative NT-related maps were derived from normative positron-emission tomography (PET) data of various neurotransmitter systems provided by Hansen et al.^19^. They include receptor and transporter density information for the serotonergic (5-HT_1a_, 5-HT_1b_, 5-HT_2a_, 5-HT_4_, 5-HT_6_, 5-HTT), dopaminergic (D1, D2, DAT), cholinergic (α_4_β_2_, VAChT, M1), glutamatergic (mGLuR_5_, NMDA), GABAergic (GABA_A_), and noradrenergic (NAT) NT systems. Each NT map was coregistered to an averaged structural connectome derived from 422 Human Connectome Project (HCP) – Aging participants, comprising 2 million streamlines (SL). For each SL, the NT-specific weight (SL_NTW_) was calculated by multiplying the respective receptor and transporter densities (*ρ*) of the NT system (*k*) at its two endpoints:

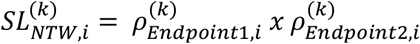

By normalizing each SL_NTW_ value to the total sum of (*N*) SL weights within the corresponding NT map (*k*), comparability across NT systems was ensured:

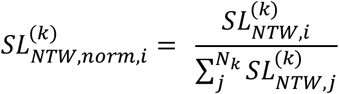

The resulting NT-related connectivity maps in MNI are publicly available via GitHub (https://github.com/phjkoch/NTDisconn). The lesion-related disruption within each NT-related system was quantified by integrating the binarized lesion masks into the NT-related connectivity maps and by identifying all streamlines intersecting the lesion area. By summing the SL_NTW_ values of all affected streamlines, an estimate of NT-specific network damage ranging from 0 (no disruption) to 1 (complete disruption of all NT-weighted streamlines) was obtained. Additionally, the total number of disconnected streamlines, regardless of their NT assignment, was computed to derive a global estimate of network damage (Disc-SL).

### Statistical analysis

Statistical analyses were performed using R version 4.4.2. To examine the relationship between damage in NT-related networks and language performance, a Partial Least Squares (PLS) analysis was conducted to select variables and identify the NT systems most strongly associated with behavioral variance. PLS regression is a dimension-reduction method that is particularly suited for analyzing multicollinear predictors, as is the case for NT-related damage scores, by projecting them onto orthogonal latent components that maximize covariance with the outcome^20^. The resulting NT systems and their corresponding damage values were subsequently entered into multiple linear regression models for further interpretability. PLS regression was implemented using the *plsr* (R package *pls*) function and included all neurotransmitter-related network damage scores as well as Disc-SL. For each predictor, Variable Importance in Projection (VIP) scores were computed across three latent components, which were retained to capture distributed covariance patterns while maintaining stable variable weights^20,21^. All predictors with VIP scores > 1 were considered informative^22^ and selected for subsequent analysis.

For each informative predictor, a separate multiple linear regression model was fitted with language performance as the dependent variable. All models were adjusted for age, sex, time post stroke, and stroke volume. In the ARC dataset, time post-stroke was log-transformed to reduce skewness. Stroke volume was cube root-transformed to reduce skewness, and within each NT-specific model, volume was linearly residualized against the respective NT-related damage score, such that the residual volume term captured variance in lesion size independent of NT-related network damage^23^. This approach mitigates collinearity while retaining NT-related damage as the primary predictor, in line with previous studies^23,24^. The corresponding base model for each cohort included only the covariates, with cube root-transformed stroke volume entered without residualization. In these NT-related models, the association between NT-specific damage and language performance was quantified using two-sided t-tests on the regression coefficients. To account for multiple comparisons, resulting *P*-values were corrected using the false discovery rate (FDR) procedure within each cohort. Additionally, improvement in explanatory power beyond the covariate-only base model was assessed by comparing Akaike Information Criterion (AIC) values, with ΔAIC defined as AIC_Baseline_ - AIC_Neurotransmitter_. Positive ΔAIC values indicate an improved fit of the NT-related model, and values greater than 2 were interpreted as a relevant gain in model performance. By comparing results across cohorts and language measures to identify convergent effects, predictors exhibiting concordant effect directions and statistically supported associations were regarded as robust neurochemical markers of post-stroke language impairment.

In a complementary analysis, lesion load was also quantified within language-associated regions^25^ defined in the Brainnetome Atlas^26^. Regional lesion loads were calculated as the proportion of lesioned voxels per region relative to the total region volume. Regions with critically skewed data distribution (median = 0 or -1.3 > skew > 1.3)^27^ were excluded. These regional measures were entered into a separate PLS regression model following the same procedure as for NT-related damage, with language performance as the outcome variable. Regions with VIP > 1 were identified as informative and subsequently entered into multiple linear regression models adjusted for the same covariates. Improvement in model fit relative to the baseline model was again quantified using ΔAIC, and model performance was compared with that of the winning NT-based models to assess the relative explanatory contribution of NT-related versus region-based structural damage.

## Results

Data from 44 participants of the WSC and 226 participants of the ARC with a left-hemispheric supratentorial stroke were included in the analysis. Please see the Supplementary Material for a flowchart of the dataset composition in both cohorts. Baseline demographic and clinical characteristics for both cohorts are presented in Table 1. The two cohorts were comparable with respect to age and sex distribution. However, they exhibited distinct profiles regarding stroke characteristics: The WSC represents the acute phase (median of 12 days post stroke) with generally smaller lesions (mean 21.8 mL), whereas the ARC represents the chronic phase (median of 654 days post stroke) of post-stroke recovery with substantially larger lesion volumes (mean 123.3 mL). Figure 1 illustrates the voxel-wise overlap of stroke lesions across all participants for both cohorts.

**Tab. 1.** Demographic and clinical data of the included participants. Systematic overview of demographic and clinical characteristics of the Washington Stroke Cohort (WSC) and Aphasia Recovery Cohort (ARC). Values are presented as mean ± standard deviation or median with corresponding interquartile range (IQR), depending on the skewness of the variables. The language factor score, used in the WSC is scaled from 0 to 100, with higher values indicating better language performance.

| Variable | Washington Stroke Cohort | Aphasia Recovery Cohort |
| --- | --- | --- |
|  | N = 44 | N=226 |
| Age, years (mean, SD) | 54.2 (12.3) | 57.8 (11.2) |
| Sex, Female/Male | 20/24 | 88/138 |
| Image acquisition, days post stroke (median, IQR) | 12 (10-14) | 654 (358-1455) |
| Behavioral assessment, days post stroke (median, IQR) | 12 (10-14) | 721 (403-1765) |
| Stroke Volume (mL, mean, SD) | 21.8 (26.3) | 123.3 (92.8) |
| Language factor acute (median, IQR) | 85.6 (63.3-94.5) |  |
| WAB-AQ (median, IQR) |  | 67.1 (39.2-89.6) |

**Figure 1.**
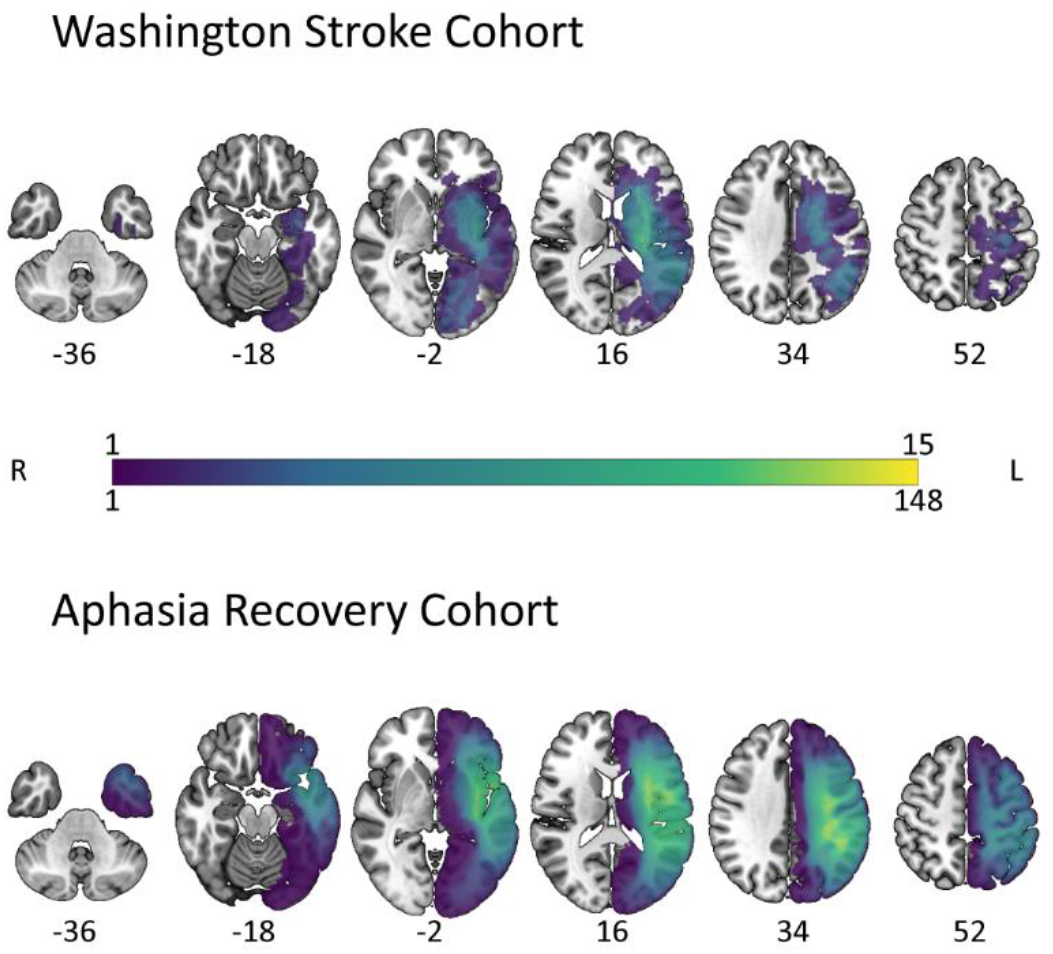
Lesion heatmap. Lesion distribution of the WSC (N = 44) and ARC (N = 226) cohort, displayed in standard MNI space. Colors indicate the number of patients with a lesion at each voxel. The numbers below the slices correspond to the z-coordinate in millimeters. R: right, L: left.

In both cohorts, damage affected all investigated NT-related networks. Still, the overall extent and pattern of involvement differed between groups (Fig. 2). The magnitude of network damage was substantially lower in the WSC (Fig. 2.A1) than in the ARC (Fig. 2.A2), reflecting the previously noted group differences in total stroke volume. PLS regression for selection of informative NT systems revealed a convergent pattern most strongly linked to language performance in both cohorts: in the WSC, damage to GABA_A_, 5-HT_1a_, D1, 5-HT_2a_, and M1-related networks emerged as the strongest contributors (VIP > 1; Fig. 2.B1); in the ARC, 5-HT_1a_, D1, 5-HT_2a_, and 5-HTT were identified as the informative predictors (Fig. 2.B2). Notably, 5-HT_1a_, 5-HT_2a_, and D1 were robustly implicated across both cohorts, indicating a consistent NT profile that generalizes qualitatively across stroke phase and lesion characteristics.

**Figure 2.**
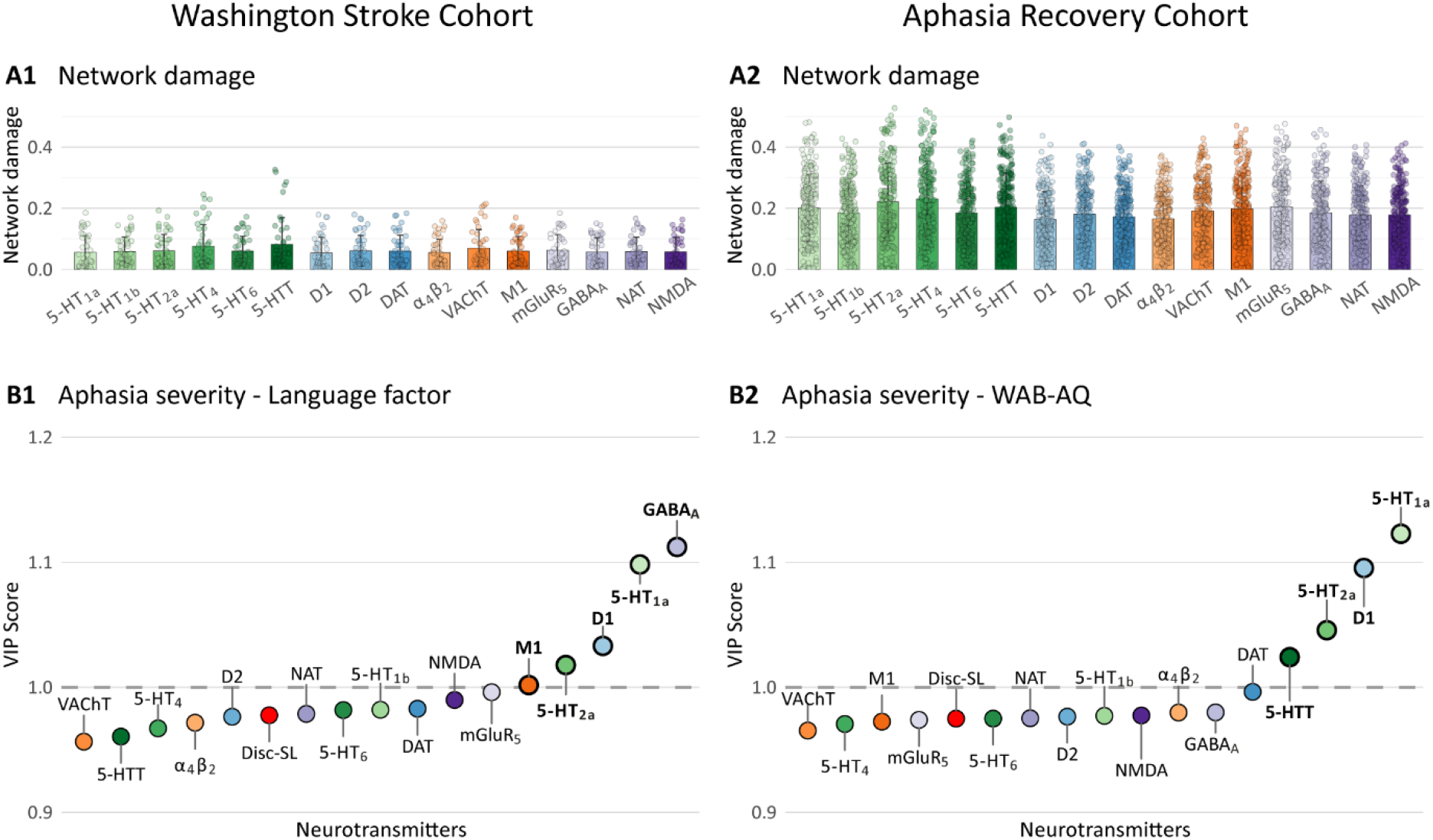
Network damage in neurotransmitter-related networks and selection of informative predictors for post-stroke language performance in WSC (acute phase) and ARC (chronic phase) Panel A visualizes the distribution of NT-related network damage of all analyzed NT systems in the Washington Stroke Cohort (A1) and Aphasia Recovery Cohort (A2). Each point represents one patient of the respective dataset. The height of the bar represents the mean NT-related network damage, and the error bar indicates the standard deviation (SD). Panel B represents the calculated Variable Importance in Projection (VIP) scores from a partial least squares (PLS) regression over 3 PLS components for each neurotransmitter in the Washington Stroke Cohort (B1) and Aphasia Recovery Cohort (B2). The PLS model was fitted to the respective language performance metric used in each dataset. The neurotransmitters are sorted by their VIP score from low to high and categorized by NT system: serotonin (green), dopamine (blue), acetylcholine (orange), and other systems (purple). Additionally, global network damage (Disc-SL) is included in this analysis and shown in red. Neurotransmitters with a VIP score > 1, highlighted by a dashed line, were considered informative. These informative predictors were selected for further analysis and are labeled in bold and highlighted with a thicker outline.

These resulting PLS-derived informative predictors were entered into adjusted multiple linear regression models for interpretability and to examine their association with the language performance after stroke (Table 2). In both cohorts, higher damage to these specific NT-related networks was consistently associated with poorer language performance (all *P*_*FDR*_ < 0.001). However, the pattern differed when assessing the added explanatory value of NT-specific damage beyond the baseline covariates (age, sex, time post stroke, and stroke volume). In the WSC, despite consistent statistical significance, the inclusion of 5-HT_1a_ (ΔAIC = 1.77) and D1 (ΔAIC = 0.96) produced only modest AIC changes and thus limited improvements in model fit. M1, GABA_A,_ and 5-HT_2a_ even yielded negative ΔAIC values, indicating that NT-related damage offered limited additional information beyond the baseline model in the acute phase. In contrast, the ARC showed substantial improvements in model fit, with the strongest improvements observed for 5-HT_1a_ (ΔAIC = 25.29), 5-HTT (ΔAIC = 24.09), and D1 (ΔAIC = 19.96).

**Table 2.** Results of multiple regression models including informative PLS predictors of language performance. For both Cohorts the most informative predictors were entered into a multiple linear regression model together with the covariates (age, sex, time post stroke, and cube root-transformed lesion volume residualized against the respective NT predictor). For interpretability, NT-related damage scores originally ranging from 0-1 were multiplied by 10, such that an increase of 1 corresponds to a 10% increase in absolute damage within the respective NT system. Reported are beta coefficients (Coeff.) for these NT-related damage scores, including 95% confidence intervals (95% CI), statistical significance (*P*_*FDR*_) of each predictor within the respective model, based on FDR-corrected p-values to account for multiple comparisons, and changes in Akaike Information Criterion (ΔAIC) compared to the baseline model including only covariates. ΔAIC was computed as AIC_Baseline_ - AIC_Neurotransmitter_, such that positive values indicate improved model fit relative to the baseline model. (^*^) indicate statistical significance. Bold neurotransmitters highlight cross-cohort consistency

| Cohort | Outcome | Neurotransmitter | Coefficient (95% CI) | $P_{FDR}$ | $\Delta AIC$ |
| --- | --- | --- | --- | --- | --- |
| Washington Stroke Cohort | Language factor | **5-HT<sub>1a</sub>** | -33.11 (-44.31 – -21.91) | < 0.001* | 1.77 |
|  |  | **D1** | -33.82 (-45.53 – -22.12) | < 0.001* | 0.96 |
|  |  | M1 | -36.24 (-49.28 – -23.19) | < 0.001* | -0.12 |
|  |  | GABA<sub>A</sub> | -37.14 (-50.79 – -23.5) | < 0.001* | -0.88 |
|  |  | 5-HT<sub>2a</sub> | -29.16 (-40.69 – -17.63) | < 0.001* | -1.47 |
| Aphasia Recovery Cohort | WAB-AQ | **5-HT<sub>1a</sub>** | -19.19 (-21.62 – -16.76) | < 0.001* | 25.29 |
|  |  | 5-HTT | -18.29 (-20.69 – -15.89) | < 0.001* | 24.09 |
|  |  | **D1** | -22.18 (-25.08 – -19.28) | < 0.001* | 19.96 |
|  |  | 5-HT<sub>2a</sub> | -15.36 (-17.57 – -13.15) | < 0.001* | 5.68 |

Taken together, 5-HT_1a_ and D1 emerged as convergent findings across both cohorts, suggesting a robust association between damage in these networks and post-stroke language performance, with greater incremental explanatory value in the chronic ARC sample (Figure 3). In the ARC, age showed a significant effect on language performance in both the baseline and the winning 5-HT_1a_ models, whereas sex showed a trend-level effect (*P* = 0.064) in the baseline model. In contrast, in the WSC, neither age nor sex showed significant effects on language performance in the baseline or winning 5-HT_1a_ models. Please see Supplementary Material for details of the winning and the baseline models.

**Figure 3.**
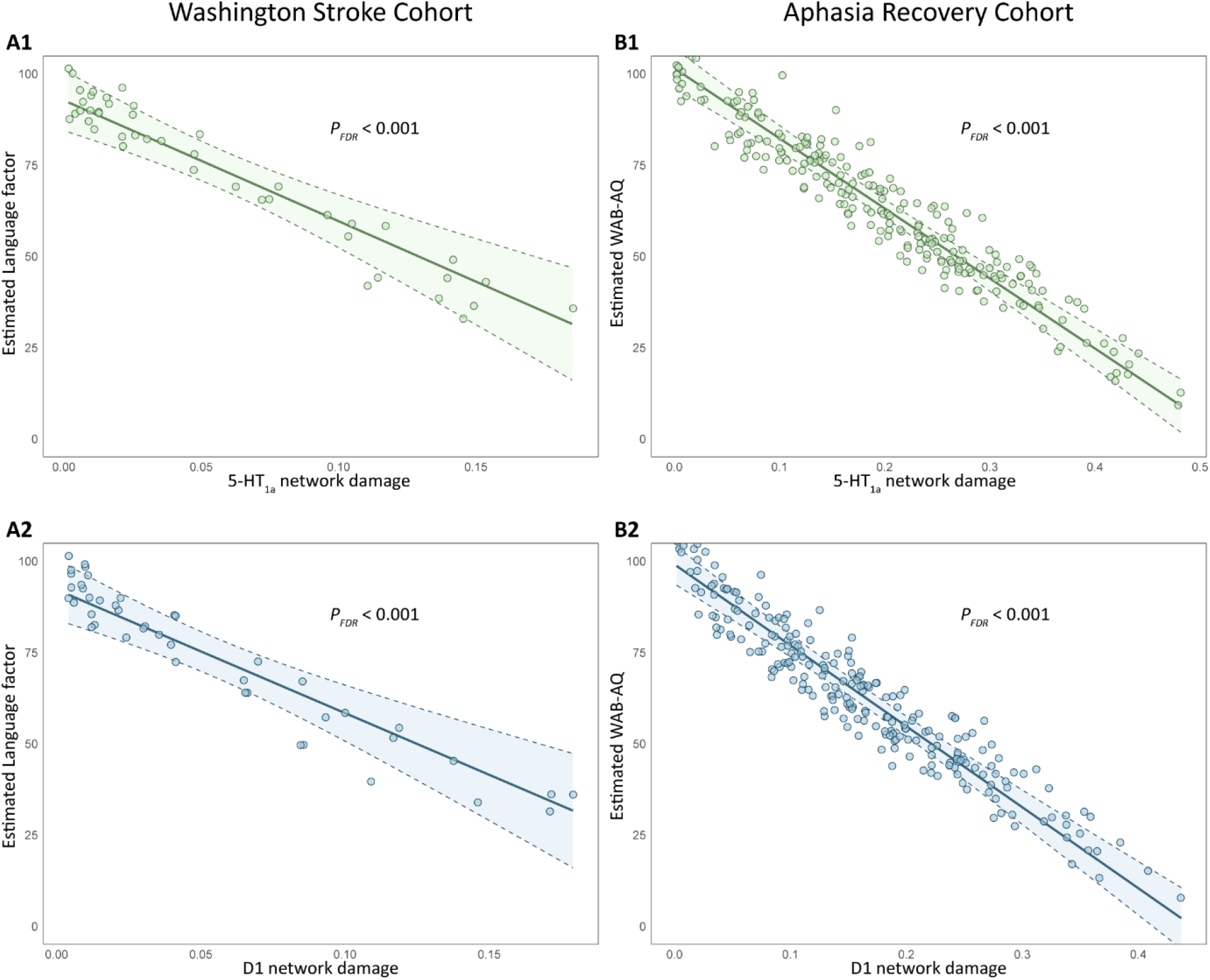
Effect Plots of 5-HT_1a_ / D1 related network damage and language performance. Effect plots illustrating the relationship between NT-related network damage and language performances in the Washington Stroke Cohort (A) and Aphasia Recovery Cohort (B). Panels show the negative association for 5-HT_1a_ (A1, B1) and D1 (A2, B2) related networks, derived from multivariable linear models adjusted for age, sex, time post-stroke, and lesion volume. Individual data points represent estimated patient performances (Language Factor for WSC; WAB-AQ for ARC), overlaid with a linear regression fit and 95% confidence intervals (shaded bands). Higher network damage is consistently associated with poorer language performance across both cohorts.

Additionally, to further probe whether NT-related network damage captures variance distinct from generic structural disconnection, a model including Disc-SL was fitted alongside the covariates, and the change in AIC relative to the base model was compared with that of the NT-related models for both cohorts. Note that Disc-SL did not reach VIP scores ≥ 1 in the PLS regression analysis, when NT data were available to be selected by the PLS model and was therefore not considered as an informative predictor in the PLS step. In the ARC, the Disc-SL model improved model fit (ΔAIC = 17.25), but was systematically outperformed by the leading NT-related models. In the WSC, the Disc-SL model failed to improve the model fit (ΔAIC = -0.49) and again remained inferior to 5-HT_1a_ and D1-based models. Regarding lesion load, in the WSC no language-associated Brainnetome region passed filtering criteria for skewed data distributions, and thus no regional lesion load analyses were conducted. In contrast, in the ARC, the tongue and larynx region in the ventral precentral and postcentral cortex (A1/2/3tonIa_L) emerged as the most informative region (ΔAIC = 3.17) but again was systematically outperformed by the leading NT-related models. These results indicate limited additional explanatory value of regional lesion load compared to NT-based measures and suggest that NT-related network damage may capture variance in post-stroke language performance beyond that attributable to structural disconnection or lesion location alone. Please see the Supplementary Material for details of the sensitivity analyses.

## Discussion

The main finding of this dual-cohort study is a robust, convergent negative association between post-stroke language performance and damage to specific serotonergic (5-HT_1a_) and dopaminergic (D1)-related brain-wide structural networks. Across two independent cohorts, the association between this neurochemical signature and language impairment remained significant after adjustment for age, sex, time post-stroke and stroke volume. However, the incremental explanatory value of these measures was more pronounced in the chronic cohort, suggesting that receptor-related network integrity may be particularly relevant for long-term language performance. This interpretation should be viewed in the context of the generally larger lesion volumes and sample size of the chronic cohort. Critically, we demonstrate that stroke-induced damage to these specific NT-related structural networks may help explain variance in language performance beyond that captured by metrics of generic structural disconnection and lesion location, highlighting their importance as innovative neurobiologically interpretable predictors of language performance after stroke.

The 5-HT_1a_ receptor exists both as an autoreceptor in the *raphe nuclei*, where it exerts inhibitory control over serotonergic firing^28,29^, and as a heteroreceptor in cortical and limbic regions^30,31^. As a heteroreceptor, it is activated by the release of serotonin in target brain regions^29^, and found in the hippocampus, septum, amygdala, and prefrontal cortex^30,31^, where it is involved in and mediates stress^29^, depression^32,33^, fear^29^, anxiety^33^, and cognitive function^29^. In the context of stroke, 5-HT_1a_ receptor agonists have been found to prevent neuronal damage after ischemic events and support neuroplasticity by increasing brain-derived neurotrophic factors (BDNF) and neurogenesis^34^. Serotonergic systems in general have long been implicated in post-stroke recovery across motor, cognitive, and language domains. However, data on the effects of serotonergic pharmacotherapy after stroke are heterogeneous. While the FLAME trial^35^ reported enhanced motor recovery in the group that was given fluoxetine and thus suggested that it could decrease stroke related disability, three larger following trials, FOCUS^36^, EFFECTS^37^ and AFFINITY^38^ failed to replicate these results and a meta-analysis of these three trials found no advantage of fluoxetine over placebo on global functional outcome or language-related domains, assessed within the Stroke Impact Scale^39^. The latter is a broad patient-reported outcome and may lack sensitivity and specificity for aphasia, underscoring that these trials were not designed to isolate effects on language recovery. In contrast, smaller aphasia-focused studies provided more encouraging evidence on the effects of SSRIs to support language performance after stroke. In a retrospective analysis, Hillis et al.^6^ found that continuous use of SSRI for the first three months after stroke was associated with a higher frequency of good improvement and a greater mean improvement in the Boston Naming Test independently of age, education, lesion volume, initial severity and depression. This effect persisted, even when key language areas were damaged. In light of the current study, one interpretation is that lesions disproportionately affecting regions embedded in 5-HT_1a_ -rich networks may compromise serotonergic modulation of plasticity and learning-related processes that support language recovery. While NT-related metrics do not directly reflect synaptic serotonin signaling, they may capture a structural correlate of regions in which serotonergic mechanisms are particularly relevant, potentially explaining why SSRI effects on language have been heterogeneous across unselected stroke cohorts. To determine whether SSRI use in acute stroke is associated with greater aphasia recovery, a prospective, multicenter, placebo-controlled randomized trial is needed. Results in this context are expected from the Protocol for Escitalopram and Language Intervention for Subacute Aphasia (ELISA)^40^ (Clinical Trials.gov ID <u>NCT03843463</u>).

D1-type receptors (D1/D5) are primarily prominent in cortical neurons^41^, and recent work suggests that D1 receptor density aligns with large-scale functional connectivity gradients in the cortex^42^. Positron emission tomography studies further indicate that dopamine release occurs in the left striatum during normal speech production and influences the lateralization of functional speech networks^43^. In general dopamine is a key contributor to motor control, motivation, learning, reward and memory processes^23^. Numerous studies evaluated the effects of dopaminergic therapy in stroke rehabilitation, yielding mixed results^23^. In fact, large randomized controlled trials challenged the concept of one-size-fits-all therapeutic approaches for stroke patients based on dopaminergic medication: The DARS^44^-trial examined the effect of Co-careldopa in addition to standard therapy, which showed no improvement in walking, physical functioning, mood, or cognition after stroke. Recently, the ESTREL^45^-trial, which tested the effect of levodopa on motor recovery in a cohort of 582 participants, showed no benefit on the Fugl-Meyer Assessment compared to placebo. For aphasia, multiple studies of bromocriptine (D2-receptor agonist) have yielded largely negative results^46,47^, whereas trials of levodopa combined with language therapy have produced mixed findings. In a prospective, randomized, placebo-controlled trial by Seniów et al., 20 patients received levodopa in addition to language therapy; those receiving levodopa showed greater improvement in verbal fluency and repetition^48^. However, these results could not be repeated by Breitenstein et al., who gave levodopa to augment high-intensity language therapy^49^, or Leeman et al., who gave it to boost an intensive computer therapy^50^. In both studies, participants improved their naming performance, but levodopa did not further enhance this effect.

Taken together, despite preclinical evidence implicating serotonergic and dopaminergic signaling in cognitive and linguistic functions, clinical translation has been hindered by inconsistent results in large pharmacological trials in stroke patients. A critical limitation of previous studies may have been the treatment of unselected, heterogeneous stroke populations without accounting for the specific neurochemical architecture of the underlying damage. Our findings address this gap at a structural level by demonstrating that damage to 5-HT_1a_ and D1-related networks is systematically associated with language performance, particularly in the chronic phase. Thus, NT-related network damage scores could serve as a novel, biologically grounded biomarker for patient stratification, paving the way for a precision-medicine approach to post-stroke aphasia rehabilitation.

### Limitations

There are several limitations to this study. First, the NT-related connectivity maps were derived from normative PET maps of aging controls and therefore do not capture individual variability in receptor and transporter density or stroke-induced neurochemical reorganization. Second, the acute WSC sample was relatively small, which may limit statistical power, particularly for multivariate approaches such as PLS. Although the pattern of associations was qualitatively consistent across cohorts, effect size estimates in the acute phase should be interpreted with caution and ideally replicated in larger samples. Third, the two cohorts differ substantially in clinical characteristics and methodology. The WSC represents an acute, general, and small stroke population with smaller lesion size and varying aphasia severity, whereas the ARC consists exclusively of chronic aphasia patients with larger lesions. This heterogeneity extends to the language testing: while the chronic cohort utilized the standardized Western Aphasia Battery, language performance in the WSC was derived from a factor analysis of multiple subscores from different standardized language assessments. However, rather than weakening the results of this study, this clinical and methodological divergence arguably underscores the robustness of our findings. Damage to serotonergic and dopaminergic networks emerged as a consistent predictor of language impairment across such distinct stroke phases, lesion characteristics, and assessment tools, which may indicate the convergent relevance of these systems that warrants further replication.

### Conclusion

In conclusion, this dual-cohort study identified a robust and convergent association between post-stroke language impairment and damage to specific serotonergic (5-HT_1a_) and dopaminergic (D1) receptor-related networks. These findings demonstrate that the vulnerability of language networks extends beyond structural disconnection to include distinct neurochemical dimensions. By providing a biologically grounded framework for characterizing individual network damage, NT-related mapping offers a promising, hypothesis-generating avenue for patient stratification and the development of targeted, precision-medicine strategies in aphasia rehabilitation, pending prospective validation in interventional studies.

## Acknowledgments

The authors acknowledge the use of an AI-based language model (ChatGPT 5.2, OpenAI) for assistance with language editing, structural refinement, and improvement of clarity. OpenEvidence was used to support literature research. All scientific content, interpretations, and conclusions were determined by the authors.

## Data availability

Publicly available datasets were analyzed in this study. The Washington Stroke Cohort data can be accessed through the Central Neuroimaging Data Archive (https://cnda.wustl.edu/app/template/Login.vm). The Aphasia Recovery Cohort data are available on OpenNeuro (https://doi.org/10.18112/openneuro.ds004884.v1.0.1). Both the NT-related connectivity maps and the code for calculating lesion-induced disconnection are publicly available via GitHub (https://github.com/phjkoch/NTDisconn).

## References

1. Flowers HL, Skoretz SA, Silver FL, Rochon E, Fang J, Flamand-Roze C, Martino R. Poststroke Aphasia Frequency, Recovery, and Outcomes: A Systematic Review and Meta-Analysis. Arch Phys Med Rehabil. 2016;97:2188–2201.e2188. doi: 10.1016/j.apmr.2016.03.006

2. Sheppard SM, Sebastian R. Diagnosing and managing post-stroke aphasia. Expert Review of Neurotherapeutics. 2021;21:221–234. doi: 10.1080/14737175.2020.1855976

3. Døli H, Andersen Helland W, Helland T, Specht K. Associations between lesion size, lesion location and aphasia in acute stroke. Aphasiology. 2021;35:745–763. doi: 10.1080/02687038.2020.1727838

4. Predictors of Poststroke Aphasia Recovery: A Systematic Review-Informed Individual Participant Data Meta-Analysis. Stroke. 2021;52:1778–1787. doi: 10.1161/strokeaha.120.031162

5. Johnson L, Basilakos A, Yourganov G, Cai B, Bonilha L, Rorden C, Fridriksson J. Progression of Aphasia Severity in the Chronic Stages of Stroke. Am J Speech Lang Pathol. 2019;28:639–649. doi: 10.1044/2018_ajslp-18-0123

6. Hillis AE, Beh YY, Sebastian R, Breining B, Tippett DC, Wright A, Saxena S, Rorden C, Bonilha L, Basilakos A, et al. Predicting recovery in acute poststroke aphasia. Ann Neurol. 2018;83:612–622. doi: 10.1002/ana.25184

7. Marebwa BK, Fridriksson J, Yourganov G, Feenaughty L, Rorden C, Bonilha L. Chronic post-stroke aphasia severity is determined by fragmentation of residual white matter networks. Sci Rep. 2017;7:8188. doi: 10.1038/s41598-017-07607-9

8. Yourganov G, Fridriksson J, Rorden C, Gleichgerrcht E, Bonilha L. Multivariate Connectome-Based Symptom Mapping in Post-Stroke Patients: Networks Supporting Language and Speech. J Neurosci. 2016;36:6668–6679. doi: 10.1523/jneurosci.4396-15.2016

9. Koch PJ, Frey BM, Backhaus W, Petersen N, Girard G, Wróbel PP, Braaß H, Bönstrup M, Kunkel Genannt Bode L, Cheng B, et al. Neurotransmitter-informed connectivity maps and their application for outcome inference after stroke. Brain. 2025. doi: 10.1093/brain/awaf185

10. Alves PN, Nozais V, Hansen JY, Corbetta M, Nachev P, Martins IP, Thiebaut de Schotten M. Neurotransmitters’ white matter mapping unveils the neurochemical fingerprints of stroke. Nat Commun. 2025;16:2555. doi: 10.1038/s41467-025-57680-2

11. Daoran Wang, Dongdong Jiang, Zhang T. Neurotransmitter Systems Underlie Structure-Function Decoupling and Recovery in Post-Stroke Aphasia. PREPRINT (Version 1) available at Research Square. 2025. doi: 10.21203/rs.3.rs-8131643/v1

12. Corbetta M, Ramsey L, Callejas A, Baldassarre A, Hacker CD, Siegel JS, Astafiev SV, Rengachary J, Zinn K, Lang CE, et al. Common behavioral clusters and subcortical anatomy in stroke. Neuron. 2015;85:927–941. doi: 10.1016/j.neuron.2015.02.027

13. Gibson M, Newman-Norlund R, Bonilha L, Fridriksson J, Hickok G, Hillis AE, den Ouden DB, Rorden C. The Aphasia Recovery Cohort, an open-source chronic stroke repository. Sci Data. 2024;11:981. doi: 10.1038/s41597-024-03819-7

14. Kertesz A. Western aphasia battery--revised. 2007. doi: 10.1037/t15168-000

15. Robb RA, Hanson DP. A software system for interactive and quantitative visualization of multidimensional biomedical images. Australas Phys Eng Sci Med. 1991;14:9–30.

16. Jenkinson M, Beckmann CF, Behrens TE, Woolrich MW, Smith SM. FSL. Neuroimage. 2012;62:782–790. doi: 10.1016/j.neuroimage.2011.09.015

17. Iglesias JE. A ready-to-use machine learning tool for symmetric multi-modality registration of brain MRI. Scientific Reports. 2023;13:6657. doi: 10.1038/s41598-023-33781-0

18. Hoffmann M, Billot B, Greve DN, Iglesias JE, Fischl B, Dalca AV. SynthMorph: Learning Contrast-Invariant Registration Without Acquired Images. IEEE Transactions on Medical Imaging. 2022;41:543–558. doi: 10.1109/TMI.2021.3116879

19. Hansen JY, Shafiei G, Markello RD, Smart K, Cox SML, Nørgaard M, Beliveau V, Wu Y, Gallezot JD, Aumont É, et al. Mapping neurotransmitter systems to the structural and functional organization of the human neocortex. Nat Neurosci. 2022;25:1569–1581. doi: 10.1038/s41593-022-01186-3

20. Wold S, Sjöström M, Eriksson L. PLS-regression: a basic tool of chemometrics. Chemometrics and Intelligent Laboratory Systems. 2001;58:109–130. doi: 10.1016/S0169-7439(01)00155-1

21. Mehmood T, Liland KH, Snipen L, Sæbø S. A review of variable selection methods in Partial Least Squares Regression. Chemometrics and Intelligent Laboratory Systems. 2012;118:62–69. doi: 10.1016/j.chemolab.2012.07.010

22. Chong I-G, Jun C-H. Performance of some variable selection methods when multicollinearity is present. Chemometrics and Intelligent Laboratory Systems. 2005;78:103–112. doi: 10.1016/j.chemolab.2004.12.011

23. Asmussen L, Frey BM, Frontzkowski LK, Wróbel PP, Grigutsch LS, Choe CU, Bönstrup M, Cheng B, Thomalla G, Quandt F, et al. Dopaminergic mesolimbic structural reserve is positively linked to better outcome after severe stroke. Brain Commun. 2024;6:fcae122. doi: 10.1093/braincomms/fcae122

24. Sadeghihassanabadi F, Frey BM, Backhaus W, Choe CU, Zittel S, Schon G, Bonstrup M, Cheng B, Thomalla G, Gerloff C, et al. Structural cerebellar reserve positively influences outcome after severe stroke. Brain Commun. 2022;4:fcac203. doi: 10.1093/braincomms/fcac203

25. Yuan B, Xie H, Wang Z, Xu Y, Zhang H, Liu J, Chen L, Li C, Tan S, Lin Z, et al. The domain-separation language network dynamics in resting state support its flexible functional segregation and integration during language and speech processing. Neuroimage. 2023;274:120132. doi: 10.1016/j.neuroimage.2023.120132

26. Fan L, Li H, Zhuo J, Zhang Y, Wang J, Chen L, Yang Z, Chu C, Xie S, Laird AR, et al. The Human Brainnetome Atlas: A New Brain Atlas Based on Connectional Architecture. Cerebral Cortex. 2016;26:3508–3526. doi: 10.1093/cercor/bhw157

27. Frontzkowski L, Fehring F, Frey BM, Wróbel PP, Reibelt A, Higgen F, Wolf S, Backhaus W, Braaß H, Koch PJ, et al. Frontoparietal Structural Network Disconnections Correlate With Outcome After a Severe Stroke. Hum Brain Mapp. 2024;45:e70060. doi: 10.1002/hbm.70060

28. Courtney NA, Ford CP. Mechanisms of 5-HT1A receptor-mediated transmission in dorsal raphe serotonin neurons. J Physiol. 2016;594:953–965. doi: 10.1113/jp271716

29. Albert PR, Vahid-Ansari F. The 5-HT1A receptor: Signaling to behavior. Biochimie. 2019;161:34–45. doi: 10.1016/j.biochi.2018.10.015

30. Pompeiano M, Palacios JM, Mengod G. Distribution and cellular localization of mRNA coding for 5-HT1A receptor in the rat brain: correlation with receptor binding. J Neurosci. 1992;12:440–453. doi: 10.1523/jneurosci.12-02-00440.1992

31. Barnes NM, Sharp T. A review of central 5-HT receptors and their function. Neuropharmacology. 1999;38:1083–1152. doi: 10.1016/S0028-3908(99)00010-6

32. Savitz J, Lucki I, Drevets WC. 5-HT1A receptor function in major depressive disorder. Progress in Neurobiology. 2009;88:17–31. doi: 10.1016/j.pneurobio.2009.01.009

33. Albert PR, Vahid-Ansari F, Luckhart C. Serotonin-prefrontal cortical circuitry in anxiety and depression phenotypes: pivotal role of pre- and post-synaptic 5-HT1A receptor expression. Front Behav Neurosci. 2014;8:199. doi: 10.3389/fnbeh.2014.00199

34. Aguiar RP, Newman-Tancredi A, Prickaerts J, Oliveira RMW. The 5-HT(1A) receptor as a serotonergic target for neuroprotection in cerebral ischemia. Prog Neuropsychopharmacol Biol Psychiatry. 2021;109:110210. doi: 10.1016/j.pnpbp.2020.110210

35. Chollet F, Tardy J, Albucher JF, Thalamas C, Berard E, Lamy C, Bejot Y, Deltour S, Jaillard A, Niclot P, et al. Fluoxetine for motor recovery after acute ischaemic stroke (FLAME): a randomised placebo-controlled trial. Lancet Neurol. 2011;10:123–130. doi: 10.1016/s1474-4422(10)70314-8

36. Effects of fluoxetine on functional outcomes after acute stroke (FOCUS): a pragmatic, double-blind, randomised, controlled trial. Lancet. 2019;393:265–274. doi: 10.1016/s0140-6736(18)32823-x

37. Safety and efficacy of fluoxetine on functional recovery after acute stroke (EFFECTS): a randomised, double-blind, placebo-controlled trial. Lancet Neurol. 2020;19:661–669. doi: 10.1016/s1474-4422(20)30219-2

38. Safety and efficacy of fluoxetine on functional outcome after acute stroke (AFFINITY): a randomised, double-blind, placebo-controlled trial. Lancet Neurol. 2020;19:651–660. doi: 10.1016/s1474-4422(20)30207-6

39. Mead G, Graham C, Lundström E, Hankey GJ, Hackett ML, Billot L, Näsman P, Forbes J, Dennis M. Individual patient data meta-analysis of the effects of fluoxetine on functional outcomes after acute stroke. Int J Stroke. 2024;19:798–808. doi: 10.1177/17474930241242628

40. Stockbridge MD, Fridriksson J, Sen S, Bonilha L, Hillis AE. Protocol for Escitalopram and Language Intervention for Subacute Aphasia (ELISA): A randomized, double blind, placebo-controlled trial. PLoS One. 2021;16:e0261474. doi: 10.1371/journal.pone.0261474

41. Civelli O, Bunzow JR, Grandy DK. Molecular diversity of the dopamine receptors. Annu Rev Pharmacol Toxicol. 1993;33:281–307. doi: 10.1146/annurev.pa.33.040193.001433

42. Pedersen R, Johansson J, Nordin K, Rieckmann A, Wåhlin A, Nyberg L, Bäckman L, Salami A. Dopamine D1-Receptor Organization Contributes to Functional Brain Architecture. J Neurosci. 2024;44. doi: 10.1523/jneurosci.0621-23.2024

43. Simonyan K, Herscovitch P, Horwitz B. Speech-induced striatal dopamine release is left lateralized and coupled to functional striatal circuits in healthy humans: a combined PET, fMRI and DTI study. Neuroimage. 2013;70:21–32. doi: 10.1016/j.neuroimage.2012.12.042

44. Ford GA, Bhakta BB, Cozens A, Cundill B, Hartley S, Holloway I, Meads D, Pearn J, Ruddock S, Sackley CM, et al. Efficacy and Mechanism Evaluation. In: Dopamine Augmented Rehabilitation in Stroke (DARS): a multicentre double-blind, randomised controlled trial of co-careldopa compared with placebo, in addition to routine NHS occupational and physical therapy, delivered early after stroke on functional recovery. Southampton (UK): NIHR Journals Library; 2019.

45. Engelter ST, Kaufmann JE, Zietz A, Luft AR, Polymeris A, Altersberger VL, Wiesner K, Wiegert M, Held JPO, Rottenberger Y, et al. Levodopa Added to Stroke Rehabilitation: The ESTREL Randomized Clinical Trial. JAMA. 2025;334:1523–1532. doi: 10.1001/jama.2025.15185

46. Zhang X, Shu B, Zhang D, Huang L, Fu Q, Du G. The Efficacy and Safety of Pharmacological Treatments for Post-stroke Aphasia. CNS Neurol Disord Drug Targets. 2018;17:509–521. doi: 10.2174/1871527317666180706143051

47. Ashtary F, Janghorbani M, Chitsaz A, Reisi M, Bahrami A. A randomized, double-blind trial of bromocriptine efficacy in nonfluent aphasia after stroke. Neurology. 2006;66:914–916. doi: 10.1212/01.wnl.0000203119.91762.0c

48. Seniów J, Litwin M, Litwin T, Leśniak M, Członkowska A. New approach to the rehabilitation of post-stroke focal cognitive syndrome: effect of levodopa combined with speech and language therapy on functional recovery from aphasia. J Neurol Sci. 2009;283:214–218. doi: 10.1016/j.jns.2009.02.336

49. Breitenstein C, Korsukewitz C, Baumgärtner A, Flöel A, Zwitserlood P, Dobel C, Knecht S. L-dopa does not add to the success of high-intensity language training in aphasia. Restor Neurol Neurosci. 2015;33:115–120. doi: 10.3233/rnn-140435

50. Leemann B, Laganaro M, Chetelat-Mabillard D, Schnider A. Crossover trial of subacute computerized aphasia therapy for anomia with the addition of either levodopa or placebo. Neurorehabil Neural Repair. 2011;25:43–47. doi: 10.1177/1545968310376938

